# The Safety, Acceptability and Efficacy of *Alena*, a Modularized CBT-based Mobile App Intervention for Social Anxiety: a Randomised Controlled Trial

**DOI:** 10.1101/2023.05.30.23288513

**Authors:** Mona M. Garvert, Stuart Linke, Tayla McCloud, Sofie S. Meyer, Sandra Sobanska, Alexander Long, Quentin J. M. Huys, Mandana Ahmadi

## Abstract

Social anxiety disorder is a debilitating mental health condition characterized by intense fear of social situations that can significantly impair daily life if left untreated. Cognitive behavioral therapy (CBT) is an effective treatment, but many patients experience slow progress, possibly due to the heterogeneity of cognitive dysfunction contributing to the maintenance of the disease that is not adequately reflected in a typical one-size-fits-all CBT approach. In addition, many patients only seek treatment late, because human therapists can themselves constitute phobic stimuli. To address these challenges, we developed *Alena,* a digital CBT program based on the Clark and Wells model of social anxiety (Clark and Wells, 1995) that was reorganized to target the four key cognitive functions associated with the maintenance of social anxiety disorder in separate therapy modules: negative beliefs, self-directed attention, rumination, and avoidance behaviors. Here, we tested the safety, acceptability and efficacy of this therapy program. In a randomized controlled trial, primary outcomes showed the app to be safe and acceptable. Secondary endpoint analyses showed that SPIN scores were significantly reduced in the treatment compared to the control group, and a larger number of participants who completed the four-week digital CBT program showed a reliable reduction in their social anxiety scores compared to a waitlist control group. Our findings suggest that targeted digital CBT without therapist involvement was safe, acceptable and showed promising signs of rapid efficacy in the treatment of social anxiety disorder.

## Introduction

Social anxiety disorder (SAD) is a severe mental health condition characterised by a persistent fear of social situations in which the individual may be scrutinised by others (American Psychiatric Association, 2013). Common scenarios that trigger social anxiety include meeting new people, speaking in public, or engaging in conversations. Individuals with social anxiety often experience a high degree of functional limitation in their everyday lives and are at increased risk for developing comorbid conditions such as mood disorders, generalized anxiety or panic disorders (Fehm et al., 2008; Kessler, 1994). SAD affects approximately 13% of people in the United States at some point in their lifetime (Kessler, 1994; Magee, 1996) and is typically a chronic condition that requires treatment for symptom improvement (Bruce et al., 2005; Reich et al., 1994).

Cognitive behavioral therapy (CBT) is the most effective treatment recommended by healthcare providers, such as the National Institute for Clinical Excellence (NICE) in the United Kingdom. CBT has demonstrated moderate to large effects on social anxiety symptoms (Acarturk et al., 2009; Mayo-Wilson et al., 2014; Powell et al., 2020). However, not all individuals respond equally well to this intervention. This is likely due to the fact that different cognitive processes contribute to the maintenance of the disorder, and individual anxiety symptoms are driven by unique cognitive profiles (Reiter et al., 2021). Clark and Wells identified four main factors underlying the maintenance of the disorder: negative beliefs, self-directed attention, anticipatory and post-event processing and the use of safety behaviors that prevent the disconfirmation of negative beliefs (Clark & Wells, 1995). While many traditional CBT programs for social anxiety address these factors through psycho-education and skills training, treatment is often not adjusted to individual cognitive profiles, resulting in time-consuming trial-and-error procedures and increasing the risk of reduced patient engagement. To improve treatment outcomes, individualized CBT programs that target specific cognitive processes may be necessary.

This problem is compounded by the fact that many individuals with symptoms of SAD never seek or receive any treatment, or only do so after many years of experiencing symptoms (Wang et al., 2005). This may be due to two main factors. Firstly, people with social anxiety symptoms may avoid seeking help due to embarrassment or fear of social scrutiny, including from a therapist who may be construed as a phobic stimulus (Lee & Stapinski, 2012). This may be compounded by concerns regarding stigmatization due to a mental illness. Secondly, traditional CBT requires regular contact with a therapist, and in many countries, there are not enough therapists available to meet demand (Davies et al., 2014). This can lead to long wait times or difficulty accessing treatment, exacerbating the problem of untreated SAD.

In response to the limited availability of trained therapists and the reluctance on the part of some patients to seek in-person treatment, several internet-based and digital versions of internet-delivered CBT (iCBT) for SAD have emerged in recent years (Andersson, 2009, 2018; Helgadóttir et al., 2014; Howell et al., 2019; Powell et al., 2020). With iCBT, patients use self-guided digital versions of the manualized CBT programme that a therapist would otherwise deliver in person. A meta-analysis of 21 randomised controlled trials testing the efficacy of these iCBT programmes found a large effect on self-reported SAD symptoms post-intervention when compared to a passive control condition, and similar effect sizes to active control conditions such as participation in an online discussion group (Andersson et al., 2012; Kampmann et al., 2016). Self-guided mobile iCBT addresses many of the barriers preventing people from accessing traditional CBT. Not having to physically attend and interact with another human in person may be particularly valuable for those with SAD. It is accessible to anyone with a smartphone and internet access, eliminating wait times and enabling users to access the programme at their convenience and without others knowing that they do so. Moreover, because it does not require any contact with a therapist, it is more cost-effective than traditional CBT. However, treatment programmes without any therapist involvement also harbour risks as some of the safeguards which can be implemented by experienced clinicians may be missing.

We developed the *Alena* app, a self-guided mobile iCBT programme specifically designed to address social anxiety disorder. The CBT programme implemented in the app is adapted from the Clark and Wells model of social anxiety (Clark & Wells, 1995). It follows a modular structure, with each module targeting a core cognitive component contributing to the maintenance of social anxiety, including negative beliefs, self-focused attention, post-event processing or rumination, and avoidance. In this study, we assessed the safety, acceptability and efficacy of the first *Alena* CBT programme. For the secondary endpoint of efficacy, we were interested both in the overall impact of the app, including limitations due to acceptability and compliance, as well as the standalone therapy in itself in ideal compliance conditions. We assessed these separately in an intention-to-treat and a per-protocol analysis.

## Methods

### Design

This was a six-week web-based parallel-group unblinded randomised controlled trial with a four-week intervention and a two-week follow-up. Participants were randomly allocated to receive CBT-based therapy for social anxiety on the *Alena* app or to a wait list control group, in a 1:1 ratio.

### Participants and recruitment

A screening study was set up on Prolific, using a self-report questionnaire hosted on Typeform. This screening study had a limit of 350 participants, to allow for ineligibility and dropout.

Participants were eligible if they were female; aged between 18 and 35 years old (inclusive); located in the UK; fluent in English; would be comfortable taking part in a study that included deception (due to the nature of the cognitive assessments); had an iPhone and daily access to an internet connection, and scored over 30 on the Social Phobia Inventory (SPIN; indicating at least moderate social anxiety; Connor et al., 2000).

Participants were excluded if they were currently undergoing any therapy for their mental health; had changed their usual mental health medication or dosage within the past eight weeks; scored eight or above on the Alcohol Use Disorders Identification Test for consumption (AUDIT-C; indicating higher risk of alcohol dependence; Reinert & Allen, 2007); scored two or above on the adapted drug questions (indicating higher risk of drug dependence) or had previously participated in scientific studies or user research undertaken by Alena.

Participants who were eligible based on their responses to this screening questionnaire were then added to the custom list of participants who were invited to the main study on Prolific. The main study had a limit of 100 participants (50 in each group). Due to a technical error, the treatment group included 52 participants.

### Procedures

All data were collected entirely online and all communication with participants took place through the Prolific platform, where participants were identified only using their Prolific ID. Participants did not provide their names or email addresses, except for one participant who contacted the study email address directly with a question about the completion date. Participants were reimbursed £5 for each of the weekly study assessments they completed, delivered via Prolific. Reimbursement for those in the intervention group was not contingent on intervention usage or adherence, but was contingent on completion of the assessment.

All participants who completed the screening study and were eligible were individually randomised to the two study arms. At the end of the baseline survey, participants were told which group they had been allocated to and given the relevant instructions. At this point, the intervention group was given the download link for the *Alena* app and their account details. Reminders were sent throughout Week 1 to participants who had not downloaded the app. Day one of the trial was defined as the date of allocation and the baseline questionnaire being available to complete. All randomised participants were invited to complete all of the questionnaires and sent up to three reminders per week, via the Prolific platform. Participants had a week to complete each questionnaire. Owing to the nature of a wait list control group, we could not blind the researcher or participants to group allocation.

For this study, the *Alena* app was only available on Apple smartphones running the iOS operating system and connected to the Internet. Participants randomised to receive the intervention were able to download and access the app free of charge using unique login details provided. Participants randomised to the waitlist control group were given access to the app for four weeks after the intervention group had completed the four-week intervention.

To explore whether any changes persisted, participants were invited for a final follow-up of efficacy measurements at week 6. At this point, the waitlist control group had access to the app for two weeks.

### Intervention

The *Alena* app (version 1.1.22; see Table 1, Figure 1) delivers a programme of CBT for social anxiety adapted for digital use. This programme is based on the Clark and Wells model of CBT for social anxiety (Clark & Wells, 1995), and is broken down into a short introductory module followed by four main modules, each targeting a particular aspect of social anxiety: negative beliefs, self-directed attention, rumination and avoidance. In the introductory module, participants self-assess their current level of anxiety, and reflect on their readiness to commit to completing the program (a motivational interviewing exercise). Each module contains between two and four exercises, which are designed to help participants practise the key therapeutic changes targeted in that specific module.

**Table 1.** Overview over the therapeutic content that participants in the intervention group had access to at different times throughout the RCT trial.

| Module | Component type | User-facing name | User-facing description |
| --- | --- | --- | --- |
| **Introduction (week 1)** | Explainer | What is social anxiety? | This chapter is an overview of social anxiety. You'll gain an insight into your own symptoms and how to recognise them. |
|  | Explainer | How does Alena work? | Learn more about Alena's unique approach, which combines evidence-based treatment with cutting-edge neuroscience assessments. |
| **Beliefs (week 1)** | Exercise | Map out your social anxiety | In this exercise you will learn how to use an evidence-based tool called the Social Anxiety Map. |
|  | Exercise | Understand your thought pattern | In this exercise, you will learn about the three types of negative thoughts which are common for social anxiety - worries, beliefs and images. |
| **Attention (week 2)** | Explainer | What is self-attention? | In this explainer, you will learn more about negative self-attention and the role it plays in keeping social anxiety going. |
|  | Exercise | Explore a new perspective | In this exercise you will work with a real situation from your life and learn how to see it from more than one perspective. |
|  | Exercise | Learn to shift your attention training | In this guided meditation you will learn how to shift your attention by focusing on different objects or sounds around you. |
|  | Exercise | Practice shifting attention | In this exercise, you will practise this new skill in a situation you might encounter in the real world - giving a speech. |
| **Avoidance (week 3)** | Explainer | What are safety behaviors? | Here you will learn more about avoidance and safety behaviors, and the role they play in social anxiety. |
|  | Exercise | Prepare for real-life practice | This exercise will help you plan an experiment to drop a safety behavior in a real-life social situation. |
|  | Exercise | Reflect on real-life practice | This reflection exercise will help you learn from your experiment and identify the most helpful next steps for you, in a non-judgemental and supportive way. |
| **Rumination (week 4)** | Explainer | What is rumination? | In this explainer, you will learn more about rumination, how it can spiral, and the role this plays in keeping social anxiety going. |
|  | Exercise | Learn to stop negative thoughts | In this exercise you will learn an evidence-based technique to help you break out of a negative thought spiral. |
|  | Exercise | Challenge your memory bias | In this exercise, you will learn a simple way to challenge any tendency you may have to remember social events in a negatively biased way. |

**Figure 1.**
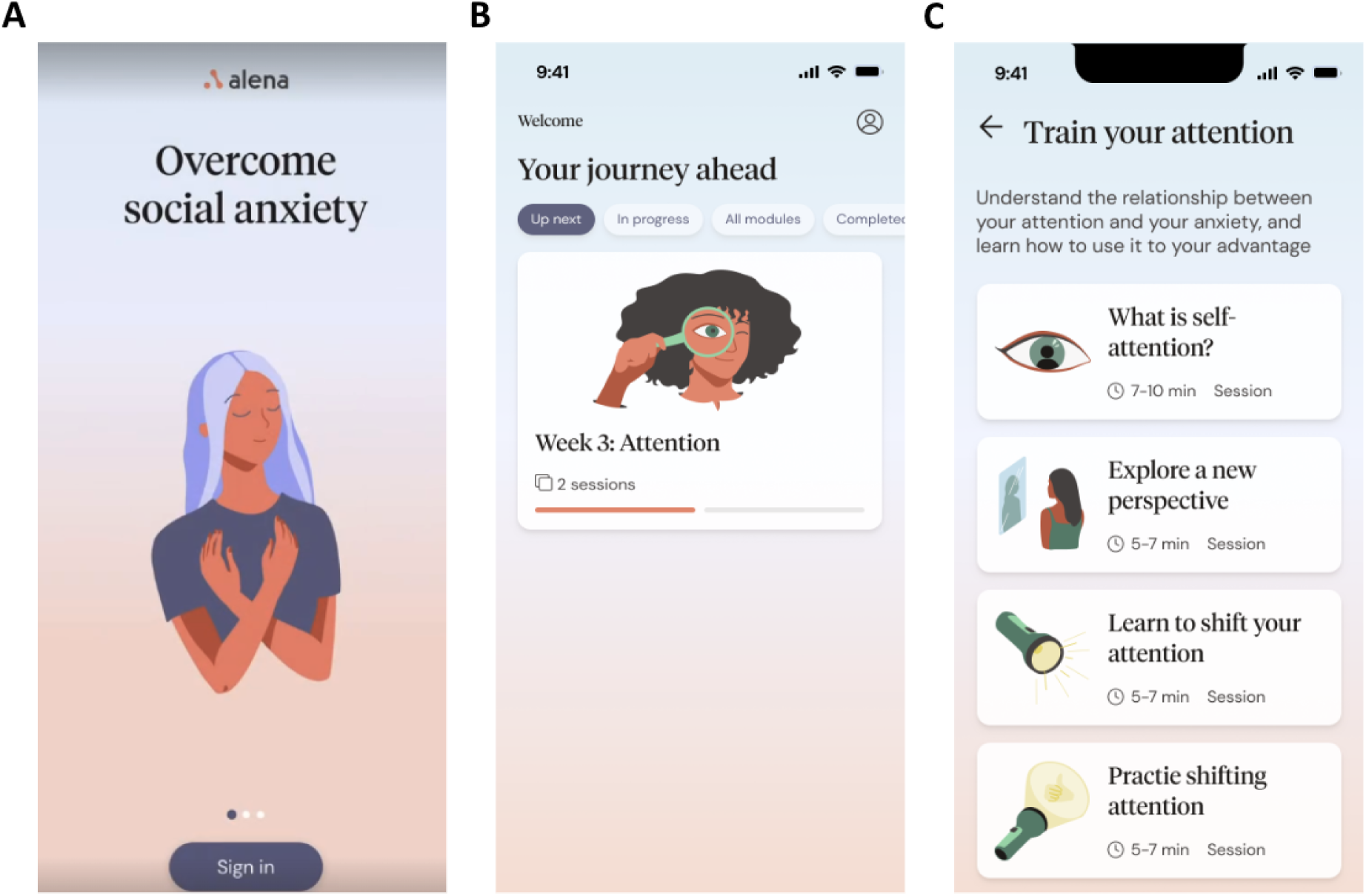
Screenshots of the *Alena* app on a smartphone. These show the onboarding page when the app is first downloaded (**A**), the home page showing the available and completed modules (**B**), and a module page showing the available exercises specifically for the attention module (**C**).

The app was developed through close collaboration between a team of scientists, clinical advisors, user researchers, designers, product managers and engineers. User feedback was obtained at a regular cadence during the development and used to continuously improve the user experience, perceived helpfulness, acceptability, design, and comprehension of user-facing content. The clinical content was based on the University College London (UCL) CBT competencies framework (Improving Access to Psychological Therapies (IAPT) Programme, 2007), implemented under the guidance of a senior clinical psychologist (SL). Exercises were predominantly text-based, with some audio. Each exercise took approximately 5-10 minutes to complete, though participants were encouraged to repeat exercises if necessary and to undertake certain activities in their own time outside of the app (for example, trying to make eye contact the next time they speak with a colleague). The app is fully automated, with no additional human involvement.

For the present study, each of the four therapy modules gradually became available on a weekly basis until all four were available in the final intervention week. Each therapy module consisted of psychoeducational content, guided psychological reflection and perspective-taking exercises, and, where appropriate, skills training exercises such as attention training, public speaking, and exposure experiments to be completed in real life. For real-life experiments, participants received guidance on how to prepare for and then reflect on the experiences (Table 1, Figure 1). Participants were also instructed to practise the skills learned in the app in their everyday life, and again supported in planning how and when to do so.

Participants were asked to complete one module per week, and were sent instructions at the beginning of each week informing them that a new module was live. The modules were labelled with their corresponding weeks to encourage participants to complete each one by the end of that week, and to complete the modules in the correct order. Participants were also encouraged to complete the exercises within each module in the order they were displayed in, and to try not to exit partway through a session. There were no prompts or reminders to use the app aside from the message at the start of each week. Data on which participants had completed each exercise was available to study researchers for the majority of (but not all) exercises, due to technical constraints.

### Demographic measures

Participant demographics (age, ethnic group, employment status and highest level of education completed) were acquired at baseline before randomization. We also asked participants whether they had ever used any mental health apps before.

### Primary Outcome Measures

Safety was assessed through a weekly questionnaire. The Intervention group was asked “Have you experienced any negative effects from using the Alena app? This could be a physical or emotional effect that you believe you have experienced as a result of using the app and/or engaging in the app therapy.” Both groups were asked: “Have you experienced any **new, serious** negative health effects in the past week? This includes having to see your GP for a new reason, going to hospital, or being otherwise very unwell in terms of your physical or mental health.” If participants responded positively to either question, they were prompted for additional details and to rate the severity of the experience.

Acceptability was also assessed through a weekly questionnaire in the intervention group. Participants were asked how satisfied they were with the app overall (Likert rating scale from very dissatisfied to very satisfied); how helpful they found the app (Likert ratings from very unhelpful to very helpful); how likely they would be to recommend the app (Likert ratings from very unlikely to very likely); how easy they found using the app (Likert ratings from very difficult to very easy); whether they got to the end of the weekly exercise (yes/no), and what got in the way of completing the exercises, with options provided.

### Secondary outcome measures

Efficacy: Specific improvement in social anxiety symptoms was measured weekly using the social phobia inventory (SPIN, Connor et al., 2000). The SPIN is an established self-report measure of the severity of social anxiety symptoms, which has been validated in various clinical and non-clinical samples (Antony et al., 2006; Connor et al., 2000). The SPIN was designed to assess the full spectrum of symptoms that characterise social anxiety, including fear, avoidance and physiological components. There are 17 items scored from 0-4, giving a total score of between 0 and 68. A score of above 19 distinguishes between socially anxious participants and controls (Antony et al., 2006; Connor et al., 2000). A reduction in SPIN score of 10 points or more from baseline indicates a reliable improvement in social anxiety (reliable change index, The National Collaborating Centre for Mental Health, 2018).

Broader improvement in functioning was measured using the Work and Social Adjustment Scale (WSAS, Mundt et al., 2002). The WSAS is a validated measure of functional impairment as a result of a particular health issue. The WSAS measures the extent to which the respondent’s problem impairs their ability to carry out day-to-day activities, such as work, home management and social leisure activities. Respondents rate each activity on a scale of 0-8 from ‘Not at all’ to ‘Very severely’, giving a total score of between 0 and 40.

### Statistical analyses

Safety: The number of adverse events were compared between the intervention and the waitlist groups using a χ^2^-test.

Acceptability: Descriptive statistics were used to characterize acceptability of the app. Specifically, we report the percentage of respondents for each item on the likert scale. Dropout was compared between the two groups at each time point using a χ^2^-test.

Efficacy: To assess the overall impact of the app, including limitations due to acceptability and compliance, we performed planned intention-to-treat linear regression analyses predicting SPIN and WSAS scores controlling for age and baseline severity for each week separately. To assess the effect of the standalone therapy itself in ideal compliance conditions, we repeated the same analysis but including only participants who completed all modules (per-protocol analysis). Descriptive statistics (mean and standard deviation) were used to report total SPIN and WSAS scores at each time point. In exploratory analyses, we additionally compared the number of participants who had experienced a reliable improvement in social anxiety by the end of the intervention (Week 4) and at follow-up (Week 6) using χ^2^ analyses. All analyses were conducted using Python version 3.8.9 (Rossum & Drake, 2010). To assess the impact of each individual therapy module on SPIN, we performed a separate linear regression analysis for each therapy module, predicting SPIN at four weeks as a function of whether or not a module was completed while controlling for age, baseline severity and total number of completed modules.

## Ethical approval

This research was approved by the Reading Independent Ethics Committee (study reference: AYSATOL). Participants viewed a digital information sheet and gave informed consent via online Typeform questionnaires before participating in both the screening and main studies. The authors assert that all procedures contributing to this work comply with the ethical standards of the relevant national and institutional committees on human experimentation and with the Helsinki Declaration of 1975, as revised in 2008.

## Results

### Participants

Of the 350 participants who completed the screening questionnaires, 158 (45.1%) were eligible to take part in the main study. Of these, we recruited 102 to take part in the main study. Participants completed the baseline questionnaires and tasks, and were randomly assigned to the treatment group (n = 50) or the intervention group (n = 52).

In the intervention group, two participants were not able to download the app during the onboarding process due to technical difficulties, and failed to do so later. All participants were sent questionnaires at all time points, regardless of whether they had downloaded the app or completed the previous questionnaire(s). Figure 2 shows the flow of participants through the trial.

**Figure 2.**
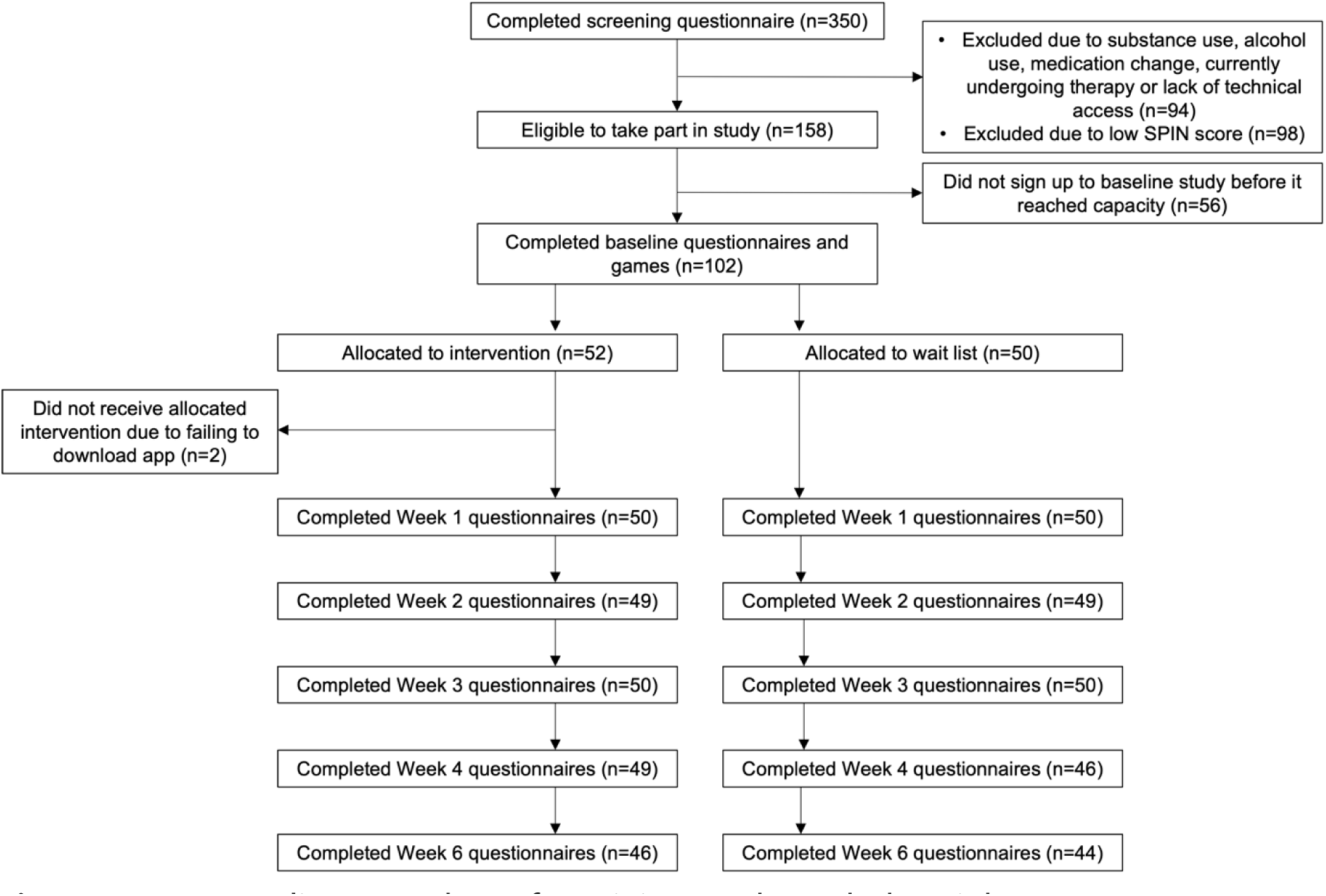
Consort diagram. Flow of participants through the trial.

### Baseline characteristics

Baseline demographic characteristics for all randomised participants are presented in Table 2. All characteristics were well balanced across groups. At baseline, the mean SPIN score was 43.5 (SD 8.38, range 31-60).

**Table 2.** Baseline demographic characteristics of study participants.

| Characteristics | Intervention<br>(n = 52) | Waitlist<br>(n = 50) | p-value |
| --- | --- | --- | --- |
| Age (years) – mean (SD) | 29.08 (4.10) | 27.46 (4.61) | 0.064 |
| Ethnicity |  |  | 0.40 |
| White/White British | 43 (82.7%) | 43 (86.0%) |  |
| Asian/Asian British, Black/Black British, Mixed/Multiple ethnic groups or other | 9 (17.3%) | 7 (24.0%) |  |
| Employment status |  |  | 0.79 |
| Full-time | 51 (61.0%) | 33 (66.0%) |  |
| Part-time | 33 (39.0%) | 7 (14.0%) |  |
| Student | 5 (9.6%) | 7 (14.0%) |  |
| Unemployed, unable to work or on temporary leave | 5 (9.6%) | 3 (6.0%) |  |
| Education |  |  | 0.38 |
| Degree(s) or other higher education | 40 (76.9%) | 31 (62.0%) |  |
| A Level or below | 12 (23.1%) | 19 (38.0%) |  |
| Has ever used mental health apps (yes) | 28 (53.9%) | 20 (40.0%) | 0.11 |

Note: Statistics are counts and percentages unless otherwise specified. SD = standard deviation.

### Safety

Four participants allocated to the intervention group (7.7%) and nine participants allocated to the waitlist group (18.0%) reported experiencing new serious, negative health effects at some point during the study (Supplementary Tables 1 and 2, χ^2^ = 1.16; p = .281). In total, fewer negative health effects were reported in the intervention group (intervention: 5 reports, waitlist: 17 reports, χ^2^ = 4.73; p = .030). None of the adverse events reported by the intervention group were rated as above moderate in severity. The other adverse events reported by the intervention group were either unrelated to intervention use (for example, a chest infection) or rated as mild or very mild.

Six adverse events were judged by participants in the intervention group as being related to using the *Alena* app. The reported events were mild to moderate in severity, and in line with what would be expected for a psychological therapy. No severe or very severe negative effects were reported from using the *Alena* app during the trial.

### Acceptability

94% of participants in the intervention group (49 of 52) and 92% in the control group (46 of 50) completed the questionnaires at Week 4, the primary endpoint and end of the intervention. At the follow-up time point two weeks later (Week 6), 88% of participants in both groups (intervention: 46 of 52, control: 44 of 50) completed the questionnaires. There was no evidence of differential dropout between the two study arms at any time point (all p > .49, χ^2^ -test).

Reported module completion decreased over time (Figure 3A), but remained above 50% in all weeks. Only four participants (7.7%) failed to complete any of the modules. Seven participants (13.5%) completed one module, eight participants (15.4%) completed two modules, and 13 participants (25.0%) completed three modules. 38.5% of participants (20 of 52) completed all four of the therapy modules (Figure 3B).

**Figure 3.**
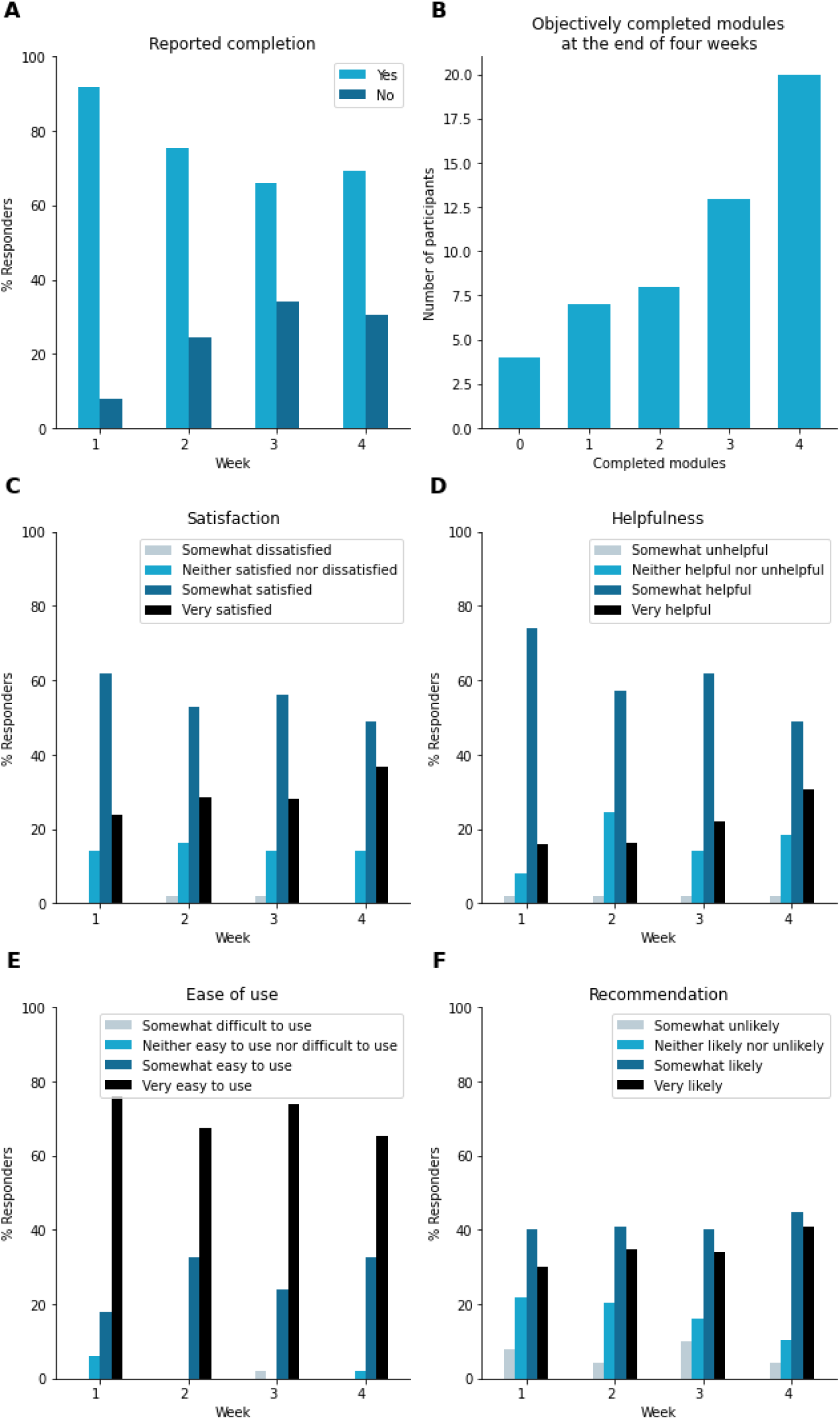
Acceptability of the intervention. A. Reported module completion by week. **B** Total number of completed modules throughout the entire study period. **C-F** Percentage of respondents reporting how satisfied they were with the app (**A**), how helpful they found the app (**B)**, how easy they found the app to use (**C**) and whether they would recommend it to someone with a similar problem (**D**).

The majority of participants in the intervention group reported feeling somewhat or very satisfied with the app (Figure 3C), finding it somewhat or very helpful (Figure 3D), and considering it somewhat or very easy to use (Figure 3E), throughout the study period. Additionally, the majority of respondents indicated that they would be somewhat or very likely to recommend the app to someone facing a similar challenge (Figure 3F).

### Efficacy

Intention-to-treat analyses showed a significant reduction in social anxiety symptoms after four weeks (primary endpoint of the intervention) in the intervention group relative to the control group (estimated mean total SPIN score difference [MD]: -5.55; 95% confidence interval [CI]: -10.54 to -0.55; p = .030; Figure 4A, Supplementary Table 3). More participants in the intervention group than in the control group experienced a reliable improvement (reduction of at least 10 points on the SPIN; intervention group (n=24/52): 46%, control group (n=9/50): 20%, χ² = 6.71; p = .009). Participants in the intervention group also reported more improvements in function (week 4 WSAS total score; MD: -2.90; 95% CI: -5.25 to -0.56; p = .016; Figure 4B, Supplementary Table 4). Hence, overall, the intervention had a beneficial effect on symptoms of social anxiety and overall function after 4 weeks.

**Figure 4.**
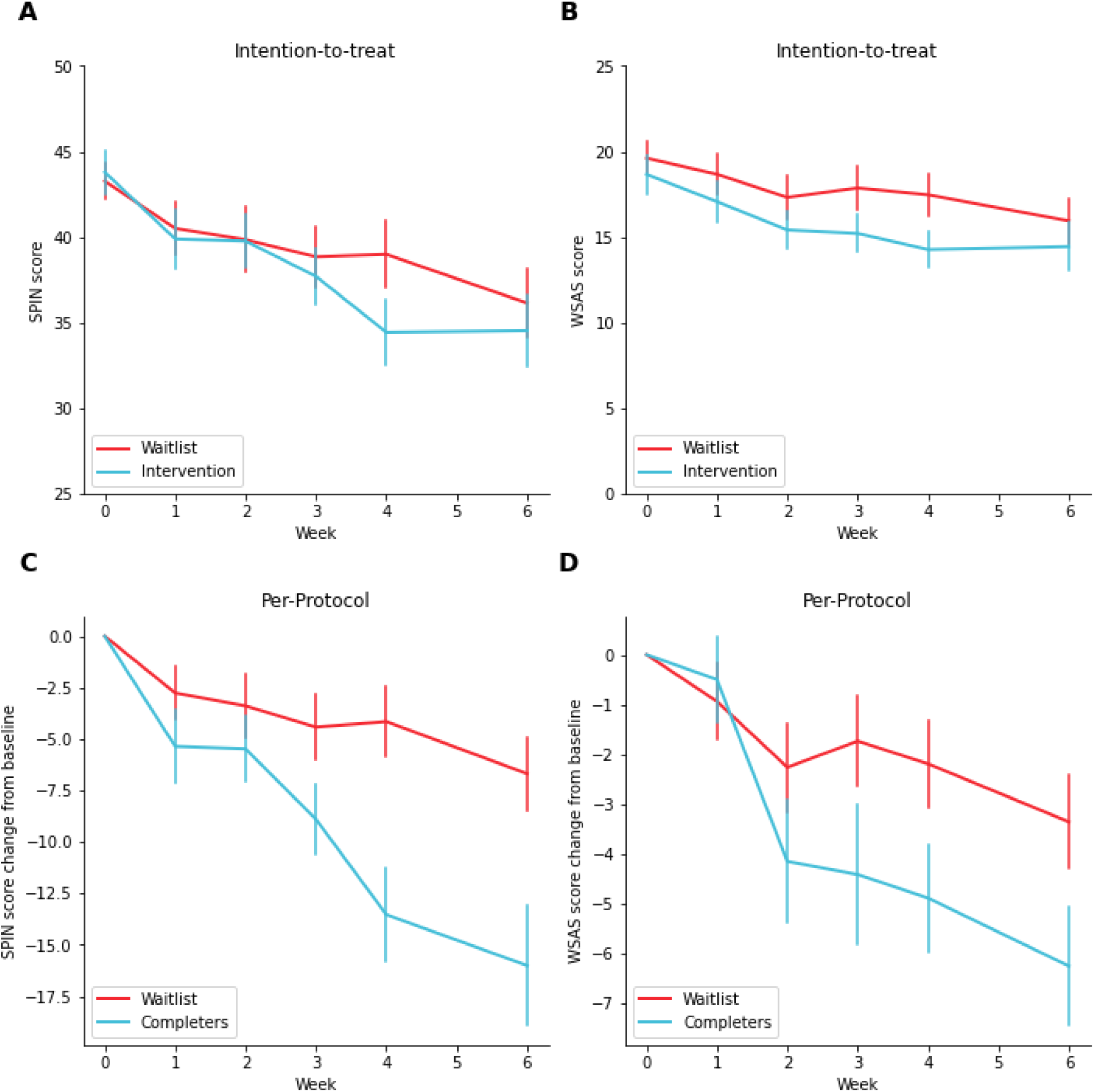
Efficacy in intention-to-treat and per-protocol analyses. A. Mean SPIN scores and **B** WSAS scores over time by study arm in the intention-to-treat sample. **C** Change in SPIN scores and **D** WSAS scores from baseline over time by study arm in the per-protocol sample. Error bars indicate standard errors of the mean.

Per-protocol analyses compared completers in the intervention group (n=20) with all of the participants in the control group (n=50). Participants in the per-protocol intervention group experienced a larger improvement than participants in the control group (SPIN total score at week 4; MD -10.83; 95% CI -17.37 to -4.29; p = .002; Figure 4C, Supplementary Table 5). A reliable change in symptoms was more frequently observed in the per-protocol intervention than in the control group (intervention group: 70% (n=14/20); control group: 20% (n=10/50): χ² = 13.71; p < .001). Functional improvement differences were also significant (WSAS total score; MD -3.64; 95% CI -6.89 to -0.39; p = .029; Figure 4D, Supplementary Table 6). Hence, most participants who completed all the modules showed an improvement in function as well as a meaningful reduction in symptoms, which was substantial at the group level.

The clinical literature suggests that exposure training is the component of CBT for social anxiety that is associated with the largest clinical benefit (Acarturk et al., 2009). To test in an exploratory analysis whether this is also true in this dataset, we compared in four separate regression analyses whether changes in SPIN at four weeks compared to baseline were related to the completion of each particular module. All regression models controlled for SPIN at baseline, age and total number of completed modules. We found that participants who completed the avoidance module in week 3 where they practised dropping a safety behavior experienced a larger change in SPIN compared to the ones who did not (estimated mean total SPIN score difference [MD]: -9.99; 95% confidence interval [CI]: -19.11 to -0.88; p = .032, Supplementary Table 7). This effect remained significant when the completion of all modules was included in the same regression model (estimated mean total SPIN score difference [MD]: --10.87; 95% confidence interval [CI]: -21.71 to -0.02; p = .050, Supplementary Table 8). Change in SPIN was not significant as a function of completing any other module (all p > .36).

### Follow-up

Improvements in symptoms were maintained at follow-up in the per-protocol analyses (MD -10.22; 95% CI -17.19 to -3.24, p = .005; reliable improvement, intervention group: 70% (n=14/20) vs control group 30% (n=15/50), χ² = 6.71; p = .009, Figure 4C, Supplementary Table 4), but not in the intention-to-treat analyses (change in total SPIN score: MD -3.38; 95% CI -8.76 to 1.99; p = .214; Figure 4A, Supplementary Table 2; reliable improvement, intervention group: 40% (n=21/52) vs control group: 30% (n=15/50), χ² = 0.79; p = .374; WSAS score improvement, group difference MD -0.91; 95% CI -3.64 to 1.82; p = .509).

## Discussion

Overall, our study suggests that the *Alena* app, a fully self-guided mobile iCBT programme for SAD based on the Clark and Wells model of social anxiety, is a safe, acceptable and efficacious tool for the treatment of social anxiety symptoms.

Most importantly, the *Alena* program for SAD was safe. This is important as the program does involve treatment steps, such as exposure, thought to necessarily induce a certain amount of discomfort, but there is no therapeutic support by experienced clinicians. Given the central importance of these components, it is critical that the therapeutic process implemented in the app would allow these components to take place in such a manner as to result in therapeutic benefit without adverse consequences. This appeared to be the case. There was no increase in adverse events severity or frequency in the intervention group compared to the control group.

Second, the *Alena* app was acceptable. We did not observe any differential dropout or bias in the participants who completed the study. Objective engagement data suggested that most participants were satisfied with the app, found it helpful, easy to use and would recommend it to others with similar problems. Many participants also used the programme consistently over time, although we did see module completion rates dropping off over time, suggesting that engagement could be improved.

Third, the Alena app was efficacious. Participants who completed the program reported significantly larger reductions in social anxiety symptoms than participants in the control group, with medium effect sizes at post-treatment and a six-week follow-up. This is consistent with previous meta-analyses that have shown the efficacy of iCBT programmes for SAD (Kampmann et al., 2016; Andrews et al., 2018).

It is important as one of the key novel aspects of the Alena app is its modular structure. The therapeutic content in the app was reorganized in an attempt to isolate and separately target specific cognitive processes with different intervention components. Because the CBT programme was broken down in this way, participants had the opportunity to focus on the cognitive components that are most relevant to their individual experience of social anxiety by repeating the corresponding exercises. This is a departure from traditional CBT one-size-fits-all programmes that may be helpful for individuals whose social anxiety symptoms are driven by specific cognitive processes. Importantly, the current results demonstrate that efficacy of iCBT is maintained when the CBT content is modularized in this way.

Changes in the Social Phobia Inventory (SPIN) are apparent specifically after completion of the avoidance module in week 3. In this module, participants learn to do exposure training, a key component of CBT for social anxiety that is often associated with the largest clinical benefit (Acarturk et al., 2009). This technique involves systematically and gradually exposing individuals to their fears in a structured manner. By repeated exposure to anxiety-provoking stimuli, participants learn corrective information about a feared stimulus (Foa & Kozak, 1986). If individuals engage with the exposure approach and persist in their efforts, they tend to gain momentum over time, even after treatment finishes, and continue to experience improvements in their symptoms.

However, our study has several limitations that should be noted. First, our sample size was relatively small. Furthermore, the sample was limited to women aged 18-35 in the UK. We chose this group as it is a relevant target demographic, since young women are the most likely to suffer from social anxiety (Weinstock, 1999). Both sample limitations mean that the results may not be generalizable to other populations, such as men or older individuals. Additionally, the study used a passive control group, which makes it difficult to draw conclusions about the specific efficacy of the Alena app compared to other interventions. Moreover, the control group did have access during follow-up which could explain the decrease in SPIN scores at follow-up in this group. Additionally, our participants were not necessarily help-seeking or previously diagnosed with SAD, which may have affected completion rates. Finally, our study only assessed the short-term efficacy of the Alena app, and it is unclear whether the benefits of the programme would be sustained over a longer period of time.

In conclusion, the findings provide preliminary support for the use of the *Alena* app as a self-guided mobile iCBT programme for SAD and motivates further work to improve engagement, and a definitive trial that is appropriately powered and longer in duration.

## Contributions

The therapeutic content was designed by SL, with support by QJMH, SS, SSM and MMG. The app implementation was led by AL, with support by SL, MMG, SSM and SS. Conceptualization and study design were led by TM, QJMH, SL, AL and MMG. Data acquisition was undertaken by TM with support by SM and MMG. Data analysis was performed by TM, MMG and QJMH. The manuscript was written by MMG and TM, with input from all authors.

## Data Availability

All data produced in the present study are available upon reasonable request to the authors.

## Acknowledgements

We would like to thank Omar Reid for the technical support in setting up this study, and the many volunteers who provided generous feedback on the Alena App during its development.

