## Supplementary Material for "The Safety, Acceptability and Efficacy of *Alena*, a Modularized CBT-based Mobile App Intervention for Social Anxiety: a Randomised Controlled Trial"

### Adverse health events

**Supplementary Table 1.** Number of reported adverse health events each week, separated by study arm.

| Week | Intervention group | Waitlist group |
|---|---|---|
| Week 1 | 1 | 3 |
| Week 2 | 2 | 3 |
| Week 3 | 1 | 4 |
| Week 4 | 0 | 4 |
| Week 6 | 1 | 3 |
| **Total no. of adverse health events** | **5** | **17** |
| **Total no. of different participants experiencing adverse events** | **4** | **9** |

**Supplementary Table 2.** Details of adverse health events reported in the two groups per week.

| Week | Intervention | Waitlist |
|---|---|---|
| 1 | ● Just had a chest infection following Covid. Follow this with GP who prescribed antibiotics. | ● Not being interested in anything<br>● I have been feeling worse than normal. I have been in a few social situations and I always feel like the odd one out.<br>● Doing physical tasks around the house that have caused slight damage to muscles and joints. |
| 2 | ● I got covid for the first time and I was hospitalised because of it. I was exhausted and in pain all week.<br>● Again chest infections being managed by antibiotics | ● I’m having severe stomach issues, symptoms of early menopause also, feeling more down and low in myself than usual<br>● Having to go up the hospital with my daughter. The place drains me mentally and takes me awhile to get back to my normal self, didn't help that we were there for hours<br>● Feeling numb |
| 3 | ● Anxious throwing up | ● Concussion<br>● Breathing problems<br>● Discussion about ADHD or Bipolar diagnosis<br>● Unable to sleep due to anxiety about new job |
| 4 | | ● Anxiety and sleep disruption caused by stress about new job<br>● Concussion recovery (weaning off painkillers)<br>● Really wanted to hurt myself. Urge<br>● Daughter fell down the stairs had to go hospital back up there again this week due to her elbow not long coming out of plaster. Stressed and full of anxiety as she can't move her arm and I'm freaking out |
| 6 | ● I had diagnostic surgery, an MRI and am awaiting more surgery for a gynae issue which has been a bit stressful. | ● Being sick effects everything i do, so feeing sorry for myself lately<br>● Extreme sleepiness/fatigue making daily activities extremely difficult. Due to chronic illness it's common for me to have fatigue but recently it has become severe and limits my already limited energy.<br>● stress |

### Adverse events related to using the app

No adverse events related to the *Alena* app were reported in weeks 1 and 4 in the intervention group.

In week 2, three mild adverse events related to using the *Alena* app were reported ('Increased anxiety from not being able to complete the task - and then guilt for not being able to.', 'The speech task made me so anxious', 'It was hard with the speeches because I've had a horrible couple weeks and my anxiety has been all over the place so I didn't feel like I could do it at the point. But I maybe could when I wasn't feeling like that').

In week 3, one very mild ('Haven't used it yet this week'), one mild ('Brought some issues to the forefront for me') and one moderate adverse event ('Forcing me to face my emotions') were reported.

### Efficacy analyses

**Supplementary table 3.** Estimated effect of Alena on SPIN scores at weeks 1-4 of the intervention and follow-up, from linear regression models.

| Week | Estimated mean difference | 95% confidence interval (CI) | p value | N |
|---|---|---|---|---|
| Week 1 | -1.56 | -5.22 to 2.11 | .401 | 100 |
| Week 2 | -1.28 | -5.30 to 2.75 | .531 | 98 |
| Week 3 | -2.15 | -6.39 to 2.08 | .315 | 100 |
| Week 4 (end of intervention) | -5.55 | -10.54 to -0.56 | .030 | 95 |
| Week 6 (follow-up) | -3.38 | -8.76 to 1.99 | .214 | 90 |

Note: Analyses include all participants with available outcome data. Each model adjusts for baseline scores and age.

**Supplementary table 4.** Estimated effect of Alena on WSAS scores at weeks 1-4 of the intervention and follow-up, from linear regression models.

| Week | Estimated mean difference | 95% confidence interval (CI) | p value | N |
|---|---|---|---|---|
| Week 1 | -0.97 | -3.02 to 1.08 | .349 | 100 |
| Week 2 | -1.54 | -3.90 to 0.82 | .197 | 98 |
| Week 3 | -2.33 | -4.79 to 0.14 | .064 | 100 |
| Week 4 (end of intervention) | -2.90 | -5.25 to -0.56 | .016 | 95 |
| Week 6 (follow-up) | -0.91 | -3.64 to 1.82 | .509 | 90 |

Note: Analyses include all participants with available outcome data. Each model adjusts for baseline scores and age.

**Supplementary table 5.** Estimated effect of Alena on SPIN scores at weeks 1-4 of the intervention and at follow-up, from linear regression models performed on per-protocol sample.

| Week | Estimated mean difference | 95% confidence interval (CI) | p value | N |
|---|---|---|---|---|
| Week 1 | -3.20 | -8.20 to 1.81 | .207 | 70 |
| Week 2 | -3.23 | -9.14 to 2.68 | .279 | 68 |
| Week 3 | -5.84 | -11.81 to 0.14 | .055 | 69 |
| Week 4 (end of intervention) | -10.83 | -17.37 to -4.29 | .002 | 65 |
| Week 6 (follow-up) | -10.22 | -17.19 to -3.24 | .005 | 63 |

Note: Analyses include all participants with available outcome data. Each model adjusts for baseline scores and age.

**Supplementary table 6.** Estimated effect of Alena on WSAS scores at weeks 1-4 of the intervention and at follow-up, from linear regression models performed on per-protocol sample.

| Week | Estimated mean difference | 95% confidence interval (CI) | p value | N |
|---|---|---|---|---|
| Week 1 | -0.46 | -3.27 to 2.34 | .741 | 70 |
| Week 2 | -2.37 | -5.84 to 1.09 | .176 | 68 |
| Week 3 | -3.52 | -7.15 to 0.11 | .057 | 69 |
| Week 4 (end of intervention) | -3.64 | -6.89 to-0.39 | .029 | 65 |
| Week 6 (follow-up) | -3.54 | -6.96 to -0.11 | .043 | 63 |

Note: Analyses include all participants with available outcome data. Each model adjusts for baseline scores and age.

**Supplementary table 7.** Estimated effect of Alena on the change in SPIN scores from week 1 to week 4 as a function of completing a particular module, from separate linear regression models performed on intention-to-treat sample.

| Module | Estimated mean difference | 95% confidence interval (CI) | p value | N |
|---|---|---|---|---|
| Module 1 | -2.54 | -16.15 to 11.06 | .708 | 49 |
| Module 2 | -3.02 | -11.88 to 5.85 | .497 | 49 |
| Module 3 | -9.99 | -19.11 to -0.88 | .032 | 49 |
| Module 4 | -5.05 | -16.13 to 6.03 | .363 | 49 |

Note: Analyses include all participants with available outcome data. Each model adjusts for baseline scores, age and total number of completed modules.

**Supplementary table 8.** Estimated effect of Alena on the change in SPIN scores from week 1 to week 4 as a function of completing a particular module, completion of all models included in the same separate linear regression model performed on intention-to-treat sample.

| Module | Estimated mean difference | 95% confidence interval (CI) | p value | N |
|---|---|---|---|---|
| Module 1 | -9.25 | --24.27 to 5.78 | .221 | 49 |
| Module 2 | -2.32 | -11.20 to 6.55 | .599 | 49 |
| Module 3 | -10.88 | -21.71 to -0.02 | .050 | 49 |
| Module 4 | -1.82 | -15.08 to 11.44 | .783 | 49 |

Note: Analyses include all participants with available outcome data. Each model adjusts for baseline scores, age and total number of completed modules.
